# Assessment of Workers’ Perception of Occupational Hazards at Nampundwe Mine in Zambia

**DOI:** 10.1101/2024.11.26.24317988

**Authors:** Anthony Phiri, Wezi Nyirenda, Enos Phiri, Maxwell Phiri, Andy Muranda, Martha Mulenga, James Manchisi

## Abstract

**Background:** The extent to which Mineworkers perceive hazards at the workplace is important to avoid Mine accidents and thus prevent damage to health, life, and property. However, Mine accidents have increased despite the large investments in health and safety training programs.

**Methods:** The study used a mixed-method approach to collect data. The study comprised 205 Workers in total, consisting of 8 Managers, 18 Supervisors, 24 Machine Operators, 80 General Miners, and 75 Contractors. These respondents were chosen by using a purposive simplifying procedure. The study used semi-structured interviews, focus group discussions, and document reviews in the collection of data.

**Results:** Study findings revealed that there was a significantly high impact of occupational health hazards on monthly income towards Contractors (p < 0.001, OR = 0.99, CI = 0.997-0.998). The study findings further reported that Workers at Nampundwe Mine have good to fair knowledge of occupational hazards. Supervisors appeared to have the highest level of awareness among the respondents, fair (38.97%), and good (54.61%). The Contractors had the lowest level of awareness and these were ranked as either fair (37, 05%) or good (18.65%).

It has also revealed that the major causes of accidents are contravening of Safety Rules (38.11%) Falling from Heights (37.52%), Trackless Vehicles (8%), Rock Falling (5.4%), Morning Shifts (4.01%) and Electricity Shocks (6.91%)

Further, the results reported that the major causes of accidents are falling from heights recorded a (37.52%) followed by trackless vehicles at (25.00%), rock falling standing at (16.06%), electricity shock (12.58%), explosion (7.10%), and least flooding at (1.74 %). Additionally, respondents are trained on how to respond to an emergency. And are very much aware of the health and safety measures and all standard operation procedures but rarely follow them when executing tasks.

**Conclusions:** Our result found Workers have good to fair knowledge of occupational hazards. Supervisors appeared to have the highest level of awareness among the respondents.

## INTRODUCTION

Globally, about 15 % of the world’s working population lives with disabilities. Many of these disability cases are found in less developed nations. Half of these disabilities result from direct injuries in the Mining Industry as opposed to the Manufacturing Industry (WHO, 2011)^[1]^.

Despite the large investments in occupational health and safety measures, Mining is still considered a dangerous industry. Studies have shown that the introduction of advanced technologies has reduced Mine Injuries by 30%. However, Mining is perceived to be a dangerous Industry compared to other Industries ^[1]^. For example, in Zambia, the worst Mine accident was recorded in 1970 in Muffler where 89 miners died as a result of flooding. In 2005 at Chambeshi Mine, 50 miners died as a result of an explosion ^[2]^. Further, 46 Miners were killed as of blast tore through an explosives factory at a Chambeshi Mine, destroying the plant ^[2]^. Globally, more than 600,000 mine workers were injured at their workplaces and more than 500,000 workers were affected by new cases of sickness caused by working in the Mine ^[3]^. In 2006, it was recorded that 3,021 non-fatal injuries occurred among coal Miners at a rate of 3.3 injuries per 100 employees who were in full employment ^[4]^. According to the Bureau of Labor Statistics ^[5]^ employee fatigue has increased the risk of accidents among workers in a Mining setup. In most workplaces, safety rules are not followed to some extent and are also not adequately explained to workers, as a result, this gives rise to accidents ^[6]^.

In Zambia, the Mining Industry employs 15% of formal workers who contribute about 8% of the Gross domestic product (GDP) and attract 84% of earnings from exports ^[7]^. A study conducted in Africa showed that there are increased rates of Mine injuries in Mine Industries when compared to those of developed nations ^[8]^. Therefore, this means that there is a need to improve the understanding of health and safety hazards in the Mining Industry. It is for this reason that the outcomes of this investigation on the perception of hazards by Mineworkers can lead to good health and safety records, increased Mine productivity, and subsequently minimize accidents, disability, and deaths.

## METHOD

### Study Site

Nampundwe Mine is located in the Central Province of Lusaka in the Sibuyunji district. (5.4999° S, 27.9308° E) It is found in the Western part of Lusaka, about 48 km from Lusaka Central Business District (CBD). The mine became operational in 1913 under the name “King Edwards Mine" ^[9]^.

The Nampundwe Mine is an underground pyrite mine that is currently owned by Konkola Copper Mine (KCM) Plc. It was previously owned by Anglo-American Cooperation, who also controlled Change Mine in Chingola, Konkola Mine in Chililabombwe, and Nampundwe pyrite mine ^[9]^. However, in 2004, the Zambian government and Vedanta Resources Limited concluded negotiations on selling 51 percent of KCM shares. Most of the shares in KCM were eventually taken by Vedanta Resources Ltd following an international bidding process led by Standard Bank and supported by the World Bank ^[10]^. The Nampundwe pyrite deposit lies on the Western flank of a synform basin with the orebody hosted within the Cheta Formation which includes massive dolomites and impure limestones, steeply dipping to the North-East. The pyrite mineralization occurs as disseminated grains, but also in continuous pyrite bands up to two meters thick^[10]^.

### Research design

According to Kothari ^[11]^, a research design informs decisions concerning a research study and the arrangement of conditions for the collection and analysis of data to combine relevance to the research. To achieve the set objective, this study employed a mixed-method approach (that is, it used both quantitative and qualitative methods) to collect data. The researcher performed data analysis in SPSS (Statisical Package for the Social Sciences). Additionally, to analyze the difference between groups of workers, the researcher used the ANOVA. To establish the different scales of knowledge whether, excellent, good, fair, or poor the researcher used the Ordinal regression. For qualitative data, the researcher made sure that all Audio files that were in focus group discussions were transcribed into computer files. After a repeat reading of the narratives, the data was later transferred into Nvivo 12 for coding.

### Sample selection

Qualitative and Quantitative data were obtained from 205 Mineworkers at Nampundwe Mine. Workers in total, consisting of 8 Managers, 18 Supervisors, 24 Machine Operators,80 General Miners and 75 Contractors.

These key informants were purposively chosen from the Mine as they have expert knowledge of the research problem under study. An interview guide was used to collect data from key informants. Purposeful sampling is a technique widely used in qualitative research for the identification and selection of information-rich cases for the most effective use of limited resources^[12]^

### Ethical considerations

Throughout the study method, social scientists have a strong moral and professional duty to act ethically ^[13]^. The first step was to obtain ethical approval from the University of Zambia’s Research Ethics Committee The participants were told about the nature of the study and their rights, which included the right to personal privacy, the right to withdraw from the research at any time without providing a reason, and the right to review and withhold interview content, all following the principles of informed consent and voluntary participation. Furthermore, there was no physical or emotional damage to the participants. The study was founded on the principles of secrecy, anonymity, and confidentiality. As a result, the participants’ identities and answers were handled with complete anonymity and privacy. To protect the confidentiality of the information, the raw data was safely stored during the study, with only the researcher and superiors having access. Finally, pseudonyms were used to protect the identities of participants and organizations when documenting the results.

### Study validity and limitations

Validity refers to the instrument’s ability to measure correctly and accurately the construct or trait that it is designed to measure ^[14]^

The researcher captured the participants at their workplace at Nampundwe Mine, this is to make sure that the research is valid. Additionally, the researcher worked closely with Supervisors, who through their experience and expertise, assisted the researcher to ensure that the research instrument was valid and not irrelevant or contradictory to the objectives of the study. To ensure validity, the questionnaire was constructed after an extensive review of relevant literature as well as instruments from similar studies conducted nationally and internationally. Validity was also ensured by aligning the flow of the questions in the instrument with the study objectives. The researcher modified the instrument according to the feedback received.

Furthermore, 12 questionnaires were first tested at Munali Nickel Mine and were revised upon taking into account the challenges encountered during the pilot study by removing ambiguities concerning the questionnaire. In order to avoid speculations made based on opinions on facts and objective information provided by the respondents. Consequently in order, to obtain empirical data, the researcher requested the respondents to provide specific indicators supporting statements, or necessary references. However, while the questionnaire might be very informative, it will at times be difficult to validate certain responses by the respondents when no data is provided in support.

Whilst limitations imply any particular study concerns potential weaknesses that are usually out of the researcher’s control and are closely associated with the chosen research design, statistical model constraints, funding constraints, or other factors. In this respect, a limitation is an ‘imposed’ restriction which is therefore essentially out of the researcher’s control ^[15]^ In this regard the Nampundwe Mine management could not avail sensitive data to the researcher. This is information in line with the number of accidents, current safety reports financial records, and many other sensitive data lastly the attainment and selection of the study respondents for the required information was difficult because workers believed that it was consuming their working time the Mine was first visited on the 20^th^ and 21^st^ August,2021. The researcher targeted six (6) FDGs however, there were challenges as workers could not all avail themselves for a focus group discussion as they were busy chasing their daily targets. More to it (4) four FDG’s where give the same information. The researcher visited the mine again on the 7^th^ and 8^th^ of May 2022. However, he could not get data for all departments of the mine hence, collected data from willing from miners workers, managers, machine operators and some contractual workers without forcing them.

## RESULTS

### Demographic characteristics

Table 1 below shows that the males were the majority at Nampundwe Mine which constituted close to 88.91% of the respondents whilst women were the minority at 11.09%. The sample size across all four categories of respondents was 4.03% Managers, 8.07% Supervisors, 20.00% Machine Operators, General Miners 50.84% and Contractors 17.06%. Furthermore, the mean age, academic education background, monthly salary, and mean years on the job are presented below in Table 1

**Table 1.**
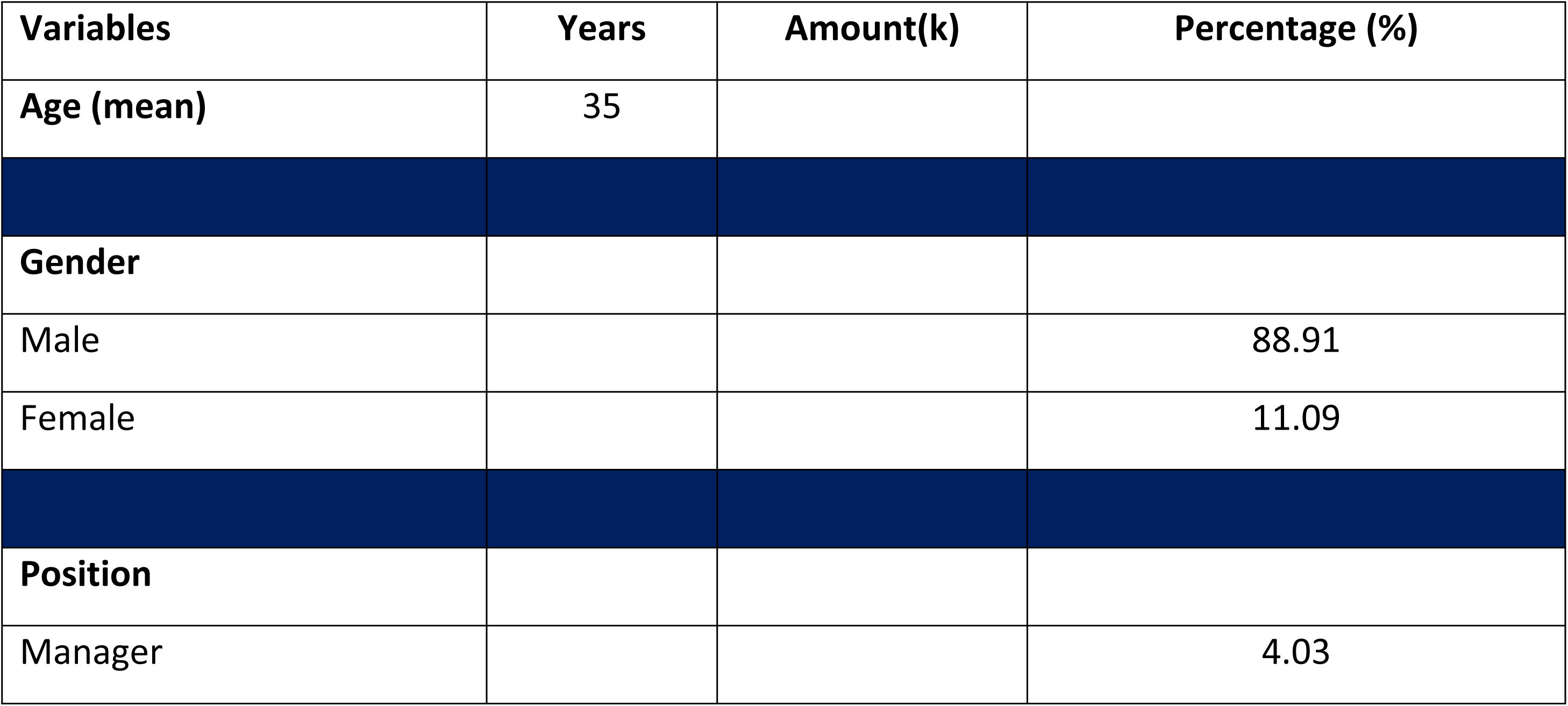
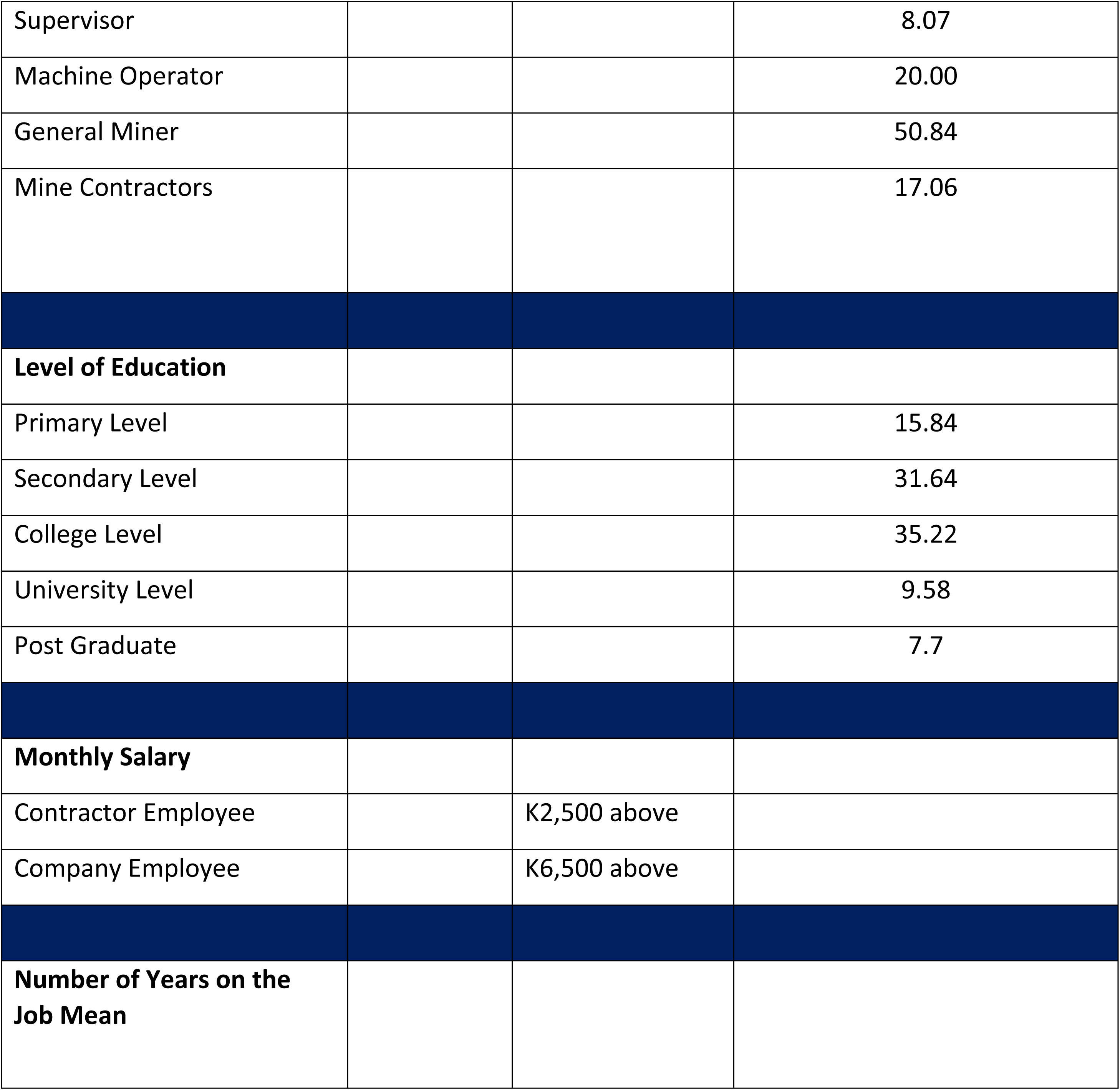
Percentage of demographic data.

| Variables | Years | Amount(k) | Percentage (%) |
| --- | --- | --- | --- |
| Age (mean) | 35 |  |  |
| Gender |  |  |  |
| Male |  |  | 88.91 |
| Female |  |  | 11.09 |
| Position |  |  |  |
| Manager |  |  | 4.03 |
| Supervisor |  |  | 8.07 |
| Machine Operator |  |  | 20.00 |
| General Miner |  |  | 50.84 |
| Mine Contractors |  |  | 17.06 |
| **Level of Education** |  |  |  |
| Primary Level |  |  | 15.84 |
| Secondary Level |  |  | 31.64 |
| College Level |  |  | 35.22 |
| University Level |  |  | 9.58 |
| Post Graduate |  |  | 7.7 |
| **Monthly Salary** |  |  |  |
| Contractor Employee |  | K2,500 above |  |
| Company Employee |  | K6,500 above |  |
| **Number of Years on the Job Mean** |  |  |  |

### Mineworkers’ Knowledge of Occupational Health and Safety at Nampundwe Mine

As shown in Table 2 below, Workers at Nampundwe Mine have good to fair knowledge of occupational hazards. Supervisors appeared to have the highest level of awareness among the respondents, fair (38.97%), and good (54.61%). The Contractors had the lowest level of awareness and these were ranked as either fair (37, 05%) good (18.65%)

**Table 2.**
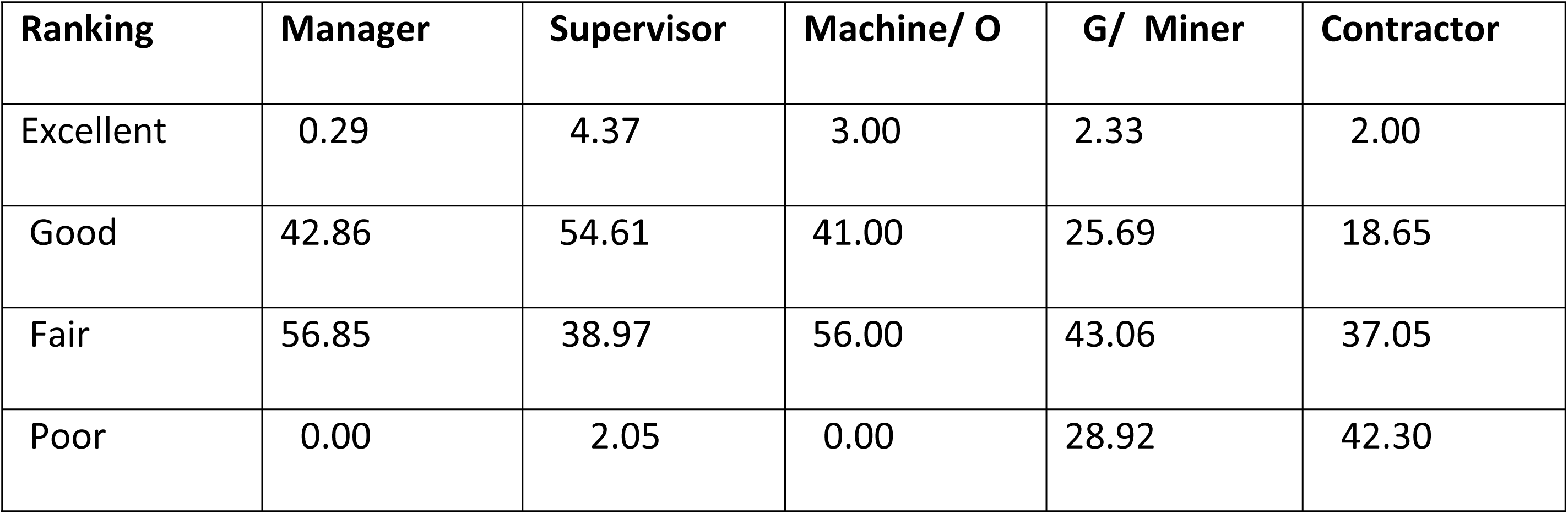
Awareness Levels of Mineworkers on the Health and Safety Hazards (%)

| **Ranking** | **Manager** | **Supervisor** | **Machine/ O** | **G/ Miner** | **Contractor** |
| --- | --- | --- | --- | --- | --- |
| Excellent | 0.29 | 4.37 | 3.00 | 2.33 | 2.00 |
| Good | 42.86 | 54.61 | 41.00 | 25.69 | 18.65 |
| Fair | 56.85 | 38.97 | 56.00 | 43.06 | 37.05 |
| Poor | 0.00 | 2.05 | 0.00 | 28.92 | 42.30 |

### Mineworker’s attitude towards the use of PPEs

As shown in Figure 2 below, Managers and Supervisors are reported to have (a 100%), attitude towards wearing Personal Protective Equipment whilst Machine Operators are reported to be 78%(160/205), General Miners 72% (147/205), and 61 %((125/205) of Contractors respectively.

**Figure 1:**
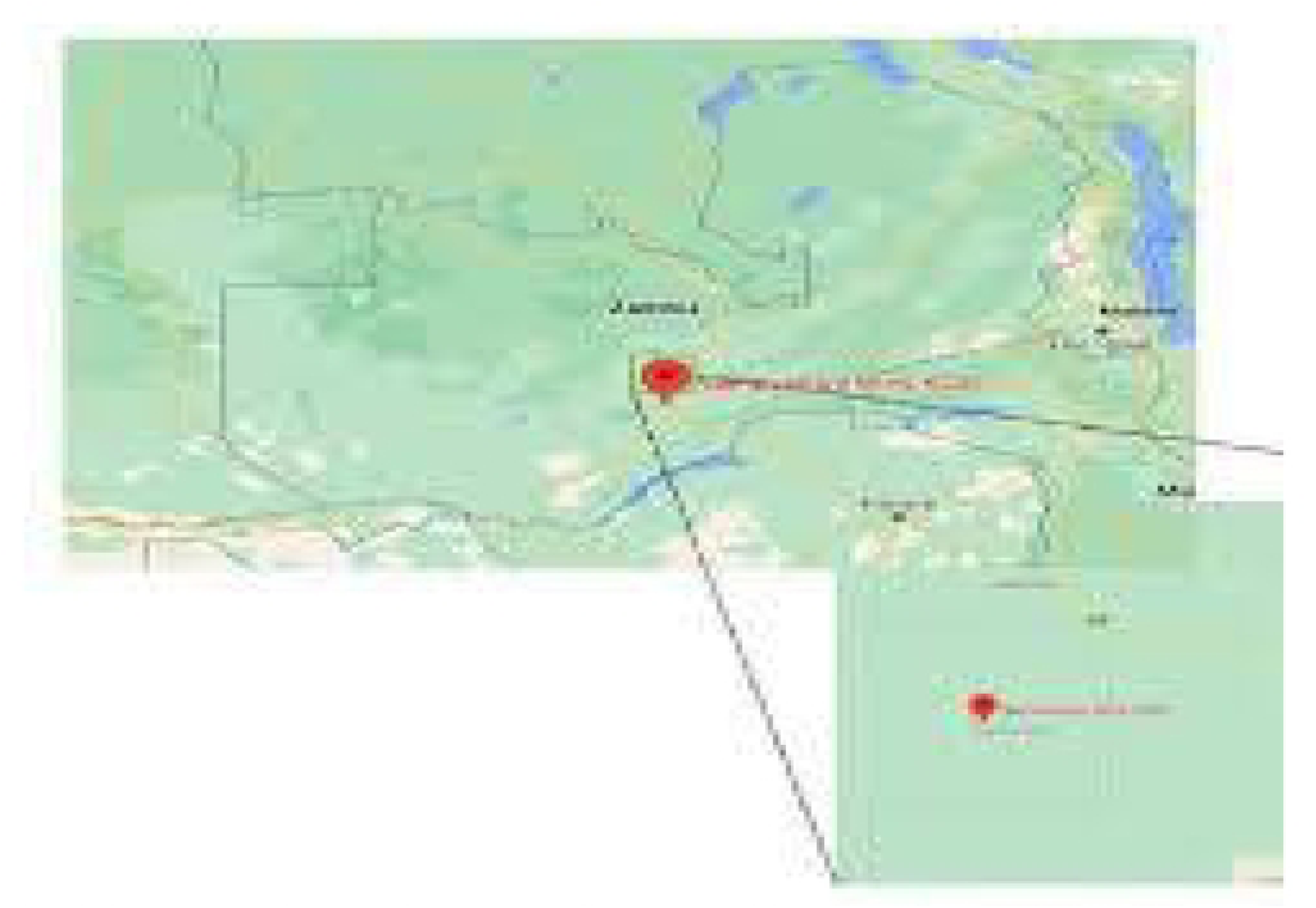
Location of Nampundwe mine in Shibuyunji District https://www.google.com/search?q=Map+of+Nampundwe+mine&rtz=1C1AVFC_enZM749ZM753&oq=Map+of+Nampundwe+mine&g

**Figure 2.**
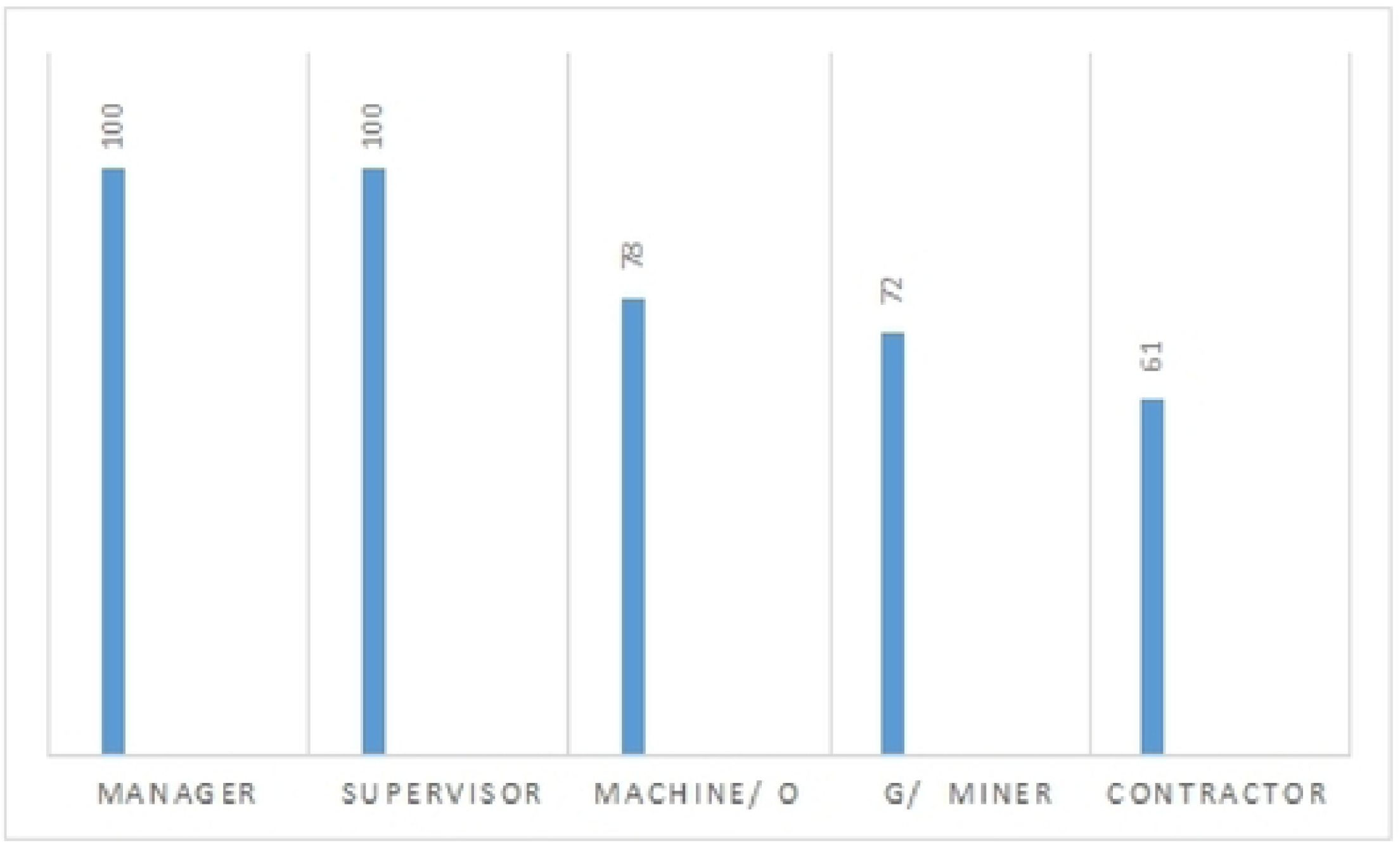
Mineworker’s attitude towards the use of PPEs (%)

### Causes of Accidents at Nampundwe Mine

As presented in Table 3, the major causes of accidents are Contravening of Safety Rules (38.11) Falling from Heights (37.52%), Trackless Vehicles (8%), Rock Falling (5.4%), Morning Shifts (4.01%) and Electricity Shocks (6.91%).

**Table 3.** Causes of accidents at Nampundwe Mine (%)

| Variables | Frequency | Percentage |
| --- | --- | --- |
| Contravening the safety rules | 12 | 38.11 |
| Morning shift | 4 | 4.01 |
| Falling from heights | 16 | 37.52 |
| Electricity shocks | 9 | 6.91 |
| Trackless Vehicle | 10 | 8 |
| Rock falling | 33 | 5.4 |

### Effectiveness of Existing Health and Safety Measures at Nampundwe Mine

As shown in Table 4 below the majority of the respondents said they had adopted the safety and health measures developed by Nampundwe Mine.

**Table 4.** Types of Health and Safety Measures Employed at Nampundwe Mine (%)

| Measure | Manager | Supervisor | Machine Operator | General Miner | Contractor |
| --- | --- | --- | --- | --- | --- |
| Safe use of workplace tools,<br>Machinery,<br>equipment | 100 | 100 | 98 | 97.56 | 96 |
| Fire safety and emergency procedures | 80 | 76.78 | 77.8 | 76 | 55 |
| First aid | 60.1 | 76.1 | 80 | 80.2 | 79 |
| Accident reporting procedure | 97 | 100 | 97 | 88.91 | 59 |
| Risk assessments | 66.1 | 60 | 57.1 | 55 | 61 |

## DISCUSSION

### Demographic characteristics

#### Age

Our study findings revealed that based on the average age of respondents of about 35 years, the mining company has a mainly youthful workforce. The majority of the workers were machine operators (at 20%) and miners (at 50.84%). As indicated in Table 1 above, the youthful age is the majority and are the ones working underground. The underground jobs are physically demanding and require great effort/endurance on the side of workers, thus making the youths most suited for the job as stated by Pransky ^[16]^. These jobs have a high risk and demand that workers are aware of safety measures to avoid or prevent mine accidents. A youthful workforce is expected to be active and would likely prevent accidents due to their quick reflexes as noted by Ilmarinen ^[17]^. Furthermore, they can easily adopt new technologies and safety measures that have the potential to minimize the occurrence of accidents. It is also possible that employed youths can easily be shifted to another department of the production line after injury without challenge. However, occurrences of accidents can only be minimized if they have adequate knowledge of safety measures and if they do adhere to the safety rules and regulations. The injuries sustained by individuals may have negative effects on the productivity of the individual as well as the company if such injuries are not properly handled ^[16]^

#### Gender

Table 1 indicates the gender of the respondents in the case study. It shows that males were represented by 88.91% and females were represented by 11.09%. The findings are similar to the World Bank report, which stated that mining companies rarely employ women because mining demands labor-intensive tasks, hence it demands men to be the majority.^[18]^

#### Position

Table 1 indicates the same position categories that existed among the respondents. There were 5 categories which were found among the respondents, namely: Manager (4.03%), Supervisor (8.07%), Machine Operator (20%), General Miner (50.84%), and Contractor (17.06%). The results findings for the respondents show that the majority of positions were Machine Operators (20%) and General Miners (50.84%)

#### Level of Education

The study findings revealed that 52.5%, (35.22%, college level, 9.58% university level, 7.7% postgraduate) Table 1, which was more than half of the respondents had tertiary education. This would suggest that the awareness levels of health and safety of hazards would be perceived with the highest comprehension. The 15.84% and 31.64 % were for primary and secondary education levels respectively. These also had an education that would appreciate awareness of hazards. Therefore, it would be ideal to state that Nampundwe Mine has an educated population that has had formal and informal education which would be an added advantage to the understanding of the awareness levels since formal education outnumbered those with informal education

#### Monthly Salary

The findings stipulate monthly earnings of a Contractor employee are in the range of K2,500 and above whilst the salaries of a Company employee are in the range of K 6,500 and above. The findings show a lack of motivation among Contractor employees that have been attributed to their low-income earnings which have caused them to breach following safety rules and regulations as stipulated by the company. This is demonstrated by comparing contractor and company employees in terms of income (Table 1). The results showed that employees with less income were less motivated and hence breached following rules and regulations compared to those with high-income earnings. As one of the Contractors stated:

> *To be honest we Contractors are very much demotivated hence we cannot follow the morning briefings on the so-called standard operational procedures which would not benefit us but would benefit more to the company …….How do you expect a Contractor employee who is getting K2,500 compared to a full-time employee who is earning K6,500 but performing the same job to follow the heavily imposed operational procedures?*

> *(Contractor; 10)*.

#### Number of Years on the Job

The study finding shows that Mineworkers had has 3 years of mean work experience where workers were highly exposed to training on health and safety and could avoid a significant number of accidents (Table 1) The reality on the ground from the Focused Group Discussion among the Mineworkers who stated that:

> *Mine workers with more than 2 years of work experience are highly aware of Mine hazards. This is because they have attended a number of training’s and a series of refresher training. However, there has been a violation of following safety rules and regulations*.

> *(Contractor; 01)*.

In this study’s findings, the demographic factors were also used for the assessment of whether there was a statistical relationship between any of them and the item that was being investigated in the questionnaire.

With an average work experience of 3 years, a Miner is expected to be familiar with almost all basic safety rules and regulations. Additionally, it is possible that they might have experienced accidents at work. For instance, Mineworkers with personal experiences of accidents at work tend to act to forestall any recurrence of accidents ^[19]^.

According to these results, workers at Nampundwe Mine have good to fair knowledge of occupational hazards. These results are corroborated in Manuele’s study ^[20]^ which reported that workers in the mining industry had reasonable knowledge of occupational hazards. These results, however, are again corroborated by Bahn’s study ^[21]^ which was completed amongst Mineworkers in Australia and also reported that the workers had good knowledge of occupational hazards.

### Mineworkers’ Knowledge of Occupational Health and Safety at Nampundwe Mine

It was also observed that Mine Managers had less knowledge level about Mine hazards, compared to Supervisors. Managers good (42.86%), fair (56.85%), and Supervisor good (54.61%) fair (38.97%). Mine Managers have the overall responsibility to directly coordinate all the duties of Supervisors and General Mineworkers. Consequently, their limited knowledge of Mine hazards entails that they are not likely to act to forestall any Mine accidents as they delegated their duties to the Supervisors. Furthermore, they are unlikely to proactively take preventive measures to minimize the risks of Mine hazards when reported by Supervisors.

It should be noted that Miners who had good or excellent awareness of the mine hazards are likely to act proactively to prevent accidents. Generally, Supervisors were more aware (56.85%) good and (4.37%) excellent about mine hazards than Managers (42.86%) good and (0.29%) excellent. Whilst Machine Operators were (41.00%) good and (3.00%) excellent, General Miners were (25.69%) good (2.33%) excellent, and Contractors (18.65%) good (2.00%) excellent. The regular interactions between Supervisors and Miners may explain the Supervisors’ better hazard awareness than Managers. Prior work experience at the mine may also account for the observed highly distinguished level of perception of hazards by Supervisors than Miners. It has been also observed from the findings that Contractors reported having very low knowledge levels of occupation hazards. This is a result of, a lack of motivation among contractor employees could have been attributed to the low-income earnings which might have affected their zeal to perform efficiently and effectively. This is demonstrated by comparing contractor and company employees in terms of income and accident records (Table 1). The results showed that employees with less income were very much demotivated and were not following health and safety rules and regulations compared to those with high-income earnings. Further, it revealed that there was a significantly high impact of occupational health hazards on monthly income towards Contractors (p < 0.001, OR = 0.99, CI = 0.997-0.998). For every 0.99 odds of decrease in monthly income, the impact from de-motivation and accidents increased, an indication of reduction of monthly income resulting from high impact de-motivation and trigger to accidents and injuries (Table 5). The findings are similar to Chipere ^[22]^ who conducted a study on stress management and reduction in the copper mines in Zambia. The factors recognized in the study that contributed to neglecting health and safety rules and stress were excess workload, low monthly income, long working hours, and unreasonable performance and demand.

**Table 5.** The Impact of Occupational Health Hazards on income levels at Nampundwe Mine. The confidence intervals are indicated at 95% and the significant differences are considered at *p* < 0.05 using ordinal logistic regression.

| Income level | Odds Ratio | Confidence Intervals | *p*-value |
| --- | --- | --- | --- |
| **Contractor** | 0.99 | 0.997 – 0.998 | 0.001 |
| **General Miners** | 2.08 | 0.95 - 4.74 | 0.071 |
| **Machine operator** | 1.32 | 0.43 – 4.01 | 0.615 |
| **Manager** | 1.24 | 0.14 – 7.69 | 0.824 |

Some poor awareness levels (73.27%), see Table 2 Supervisor (2.05%) General Miner(28.92%), and Contractor(42.30%) on the health and safety of mine hazards can be attributed to workers’ low educational attainment (15.84% were primary school level), which may have made it impossible for them to follow safety instructions within the mine, especially instructions in written format. According to Bahn ^[21]^ and Weichbrodt ^[23]^, workers who are not privy to safety rules and their implications on their health are likely to adopt a laid-back attitude towards those rules. Being able to read safety instructions is important in avoiding potential injury and accidents, and so, if workers are unable to read instructions, many accidents are likely to occur. Another possible cause of the lack of awareness is inadequate training and refresher courses on safety issues. Training would likely maximize their level of knowledge and hazard awareness to avoid accidents and injuries. Additionally, well-trained workers are likely to employ all safety measures available to avert and avoid any potential danger. They are more likely to utilize all available protective gear including clothing, boots, and helmets. They are also likely to report on any compromises in terms of safety within and around the mines. The lack of knowledge on safety measures would possibly expose workers to injuries that will lead to long-term disabilities as reaffirmed by Golovina ^[24]^. Consequently, workers who adhere to all safety rules and yet suffer accidents and/or injuries from accidents that they had little control over, such as landslides, may ignore the entire safety system. Such individuals may ignore personal safety precautions such as protective boots and bump caps, knowing very well that they had very little control over previous accidents ^[23]^.

### Mineworker’s attitude towards the use of PPEs

As shown in graph 2. The study findings revealed that (100%) of Managers and (100%) of Supervisors reported having a positive attitude towards the wearing of Personal Protective Equipment and further reported that PPE helps to prevent Mine accidents. This is similar to the study conducted by Mavhunga ^[22]^ who reported that PPEs are useful to Mineworkers and protect them from Mine hazards. However, the findings reported that 78 %(160/205) of Machine Operators, 72% (147/205) of General miners and 61 %((125/205) of Contractors respectively exhibited a reduced attitude towards the use of PPE. This is in support of the findings conducted by Rosenburg and Levenstein ^[25]^ who reported that some Mineworkers demonstrated a negative attitude towards PPE and found PPE to be "frequently uncomfortable, is rarely fully protective, and is sometimes hazardous to the health of workers wearing the equipment for long periods of time. The finding has revealed that an excellent attitude toward the use of PPE by Managers and Supervisors is a result of top management being the enforcers of good health and safety measures at the Mine. A negative attitude towards the use of PPE from a few of the Machine operators (22%), General miners (28%), and Contractors (39%) is been reported to have forbidden the act of wearing the PPE for a long time. This is in contrast with ILO, ^[26]^ reports on the importance of wearing PPE. As putting on PPE helps to protect Mineworkers from unforeseen accidents, therefore, working in the Mine without wearing PPE on a regular basis may expose Miners to avoidable accidents that may lead to injuries, disabilities, and even death.

### Causes of Accidents at Nampundwe Mine

The finding shows that at Nampundwe Mine the frequency of accidents was caused by contravening the safety rules (38.11%) see (Table 3). This is in line with findings made by Flippo^[27]^ that in America four accidents were caused by human error to everyone that is due to technical causes. His findings also postulated that human causes are connected with deficiencies in individuals such as improper attitudes, carelessness, recklessness, inability to perform the job, daydreaming, alcoholism, and the use of drugs on the job. These findings were also confirmed by a study done on lost time accident areas of occurrence and their costs done by Musonda ^[28]^at Nampundwe Mine which highlighted some of the causes of accidents as lack of supervision, failure to exchange warning whistles, reporting to work under the influence of alcohol. This is followed by falling from heights (37.52%) at Nampundwe Mine there are some slippery environments that are prone to accidents. The reality on the ground from the Focused Group Discussion among the mine workers stated that:

> *There are a number of hazardous environments in our Mine, some are slippery and some environments in the mine are not all that stable……… you would find that whilst you are drilling an earth environment would fall on you*.

> *(Mine worker; 07)*.

Thirdly, the findings reported that Trackless Vehicles (8 %) as a result of roadways which are poorly maintained, maximizes driver’s exposure to accidents. Most of the underground working places are poorly illuminated, which always results in poor driving conditions and a lack of visibility. These findings are similar to the study findings by Fourie ^[22]^ on the Analyses of South African Mine safety statistics which stated that many South African Mines accidents are associated with poor roadways, poor illumination, and the use of heavy plant and machinery as one of the challenges. However, one of the responses from the Mine Managers stated that Trackless Vehicle accidents occurred due to human error resulting from alcohol consumption (for example, Trackless Vehicle drivers tend to hide and take small sachet of alcohol during working hours). Another cause of accidents reported to be Rock Falling (5.4 %), Miners are exposed to falls of rocks. The findings reported that Rockfall occurs due to high outcrops of hard, erosion-resistant rocks that become unstable for a variety of reasons. The size of the falling rock depends on the source area, bedding thickness, bedding dip and dip direction, hardness, weathering, position, and steepness of the slope. The findings are supported by Stacey ^[29]^ who stated that Rockfall accidents continue to be the main causes of fatalities in the mining industry. Furthermore, Morning Shift at (4, 01%) The study also showed accidents occurred during the Morning Shift (Table 3). It has been reported that employees work under pressure as the employer is interested in high rate copper production and increasing the profits as the sustainability of an industry depends on high profits and as such a lot of mistakes may be committed resulting in Mine accidents.

Lastly, findings reported that (6.91 %), accidents may happen due to Electric Shock. Electrical accidents happen during drilling and during maintenance work. The findings are supported by Cawley J. C. ^[30]^ who narrated that, most of my electrical accidents and fatalities were sustained during electrical maintenance and during mineral mining.

### Effectiveness of Existing Health and Safety Measures at Nampundwe Mine

As shown in (Table 4) Health and safety measures have been employed at Nampundwe Mine where Training in basic health and safety rules of hazard identification and control have been introduced. The findings stated that all employees are trained on how to respond to an emergency. For example, when an emergency alarm is triggered, everyone is expected to rush to the emergency assembly point so that the fire team or rescue team responds accordingly. The results are similar to the findings by McLaughlin ^[31]^ who stated that if employees are trained to follow basic health and safety rules accidents can be minimized.

However, not all Mineworkers follow the rules and regulations. A possible reason for the Mineworkers who do not adhere to health and safety procedures could be their lower literacy rates. Table 1 (15.84%) respondents with attained primary school. They are likely to prefer hands-on training methods than the written health and safety measures. Poplin ^[32]^, noted that almost all workers who participate in orientation and health and safety training activities before starting work are an indication that, they are likely to be better adapted to their work and subsequently avoid serious injuries that may lead to long-term disabilities

## CONCLUSION

Our study findings revealed that Mineworkers have good to fair knowledge of occupational hazards. Mine Managers had less knowledge level about mine hazards, compared to Supervisors. Managers good (42.86%), fair (56.85%), and Supervisor good (54.61%) fair (38.97%). Mine managers have the overall responsibility to directly coordinate all the duties of Supervisors. Generally, Supervisors were more aware (56.85%) good and (4.37%) excellent about mine hazards than Managers (42.86%) good and (0.14%) excellent. Whilst Machine Operators were (41.00%) good and (3.00%) excellent, General Miners were (25.69%) good (2.33%) excellent and Contractors (18.65%) good (2.00) excellent.

## Data Availability

N/A

## ACKNOWLEDGEMENT

I would hereby take this opportunity to express my sincere gratitude to all co-authors for the support throughout this research project. I am also grateful to the Management and staff at Nampundwe Mine for providing me with the opportunity to conduct this research and for their assistance in facilitating data collection. Additionally, I would like to thank the villagers in the area for their willingness to welcome and accommodate me. Their welcome have been productive to this research. I appreciate the time and effort taken by all individuals and organizations involved in this project.

